# Association between plasma phosphorylated tau-217 and cognition in Parkinson’s disease

**DOI:** 10.64898/2025.12.04.25341636

**Authors:** Trycia Kouchache, Tevy Chan, Sophie Sun, Aline Delva, Pedro Rosa-Neto, Jean-François Gagnon, Alain Dagher, Ronald B. Postuma, Madeleine Sharp

## Abstract

**Background:** It is increasingly recognized that Alzheimer’s disease (AD) co-pathology contributes to dementia in PD, but its role in earlier stages of cognitive impairment remains uncertain. This study examined whether plasma phosphorylated Tau at threonine 217 (p-tau217), a biomarker of early AD-related pathology, is associated with cognitive impairment in PD.

**Methods:** Using cross-sectional data from 167 PD patients without dementia and 63 controls of the Quebec Parkinson Network registry we examined the association between plasma p-tau217 and three measures of cognitive impairment: performance on standard neuropsychological testing, and cognitive impairment as defined by a Montreal Cognitive Assessment (MoCA) score <26 and by self-report. Glial Fibrillary Acidic Protein (GFAP) and Neurofilament Light chain (NfL) were also measured as non-specific markers of inflammation and neurodegeneration.

**Results:** No significant difference in p-tau217 level was observed between groups; GFAP and NfL were higher in PD patients, but these differences did not survive multiple comparison correction (*pFDR* > 0.08). Among PD patients, higher p-tau217 was associated with worse visuospatial function (p=0.04) and greater self-reported cognitive impairment (p=0.03), but these associations did not survive multiple comparison correction (*pFDR* > 0.08). There was no association with cognitive impairment as defined by a MoCA <26.

**Conclusion:** Plasma p-tau217 was not clearly associated with early cognitive impairment suggesting that co-morbid AD pathology is not a major contributor to early cognitive changes in this sample of PD patients without dementia. Replication of these findings and longitudinal research is needed to determine whether p-tau217 can predict progression to dementia in PD.

## Introduction

Cognitive impairment is a common and debilitating non-motor symptom of Parkinson’s disease (PD), affecting approximately 10% of patients in early stages, with a prevalence increasing to as high as 75% as the disease progresses (1–5). One notable feature is the variability in both the timing of onset within disease progression and the severity of cognitive impairment across individuals (4). Accumulating evidence suggests that co-existing Alzheimer’s disease (AD) pathology may contribute to this heterogeneity (2,6). This idea has been supported by autopsy studies demonstrating the presence of amyloid-β plaques (Aβ) and tau neurofibrillary tangles (NFT) – hallmarks of AD pathology – in some individuals with PD (7). For instance, one autopsy study found that 41.6% of PD patients with dementia (PDD) have AD co-pathology (2). Evidence from both amyloid and tau PET imaging studies generally align with autopsy studies (8–10). For instance, one recent PET study showed increased cortical tau load in PDD, and an examination of PD patients without dementia showed that increased cortical and subcortical amyloid was associated with worse cognitive performance (11,12). However, the majority of post-mortem and PET studies have focused on PDD, i.e., the later stage of PD-associated cognitive impairment, leaving open questions about the degree to which AD pathology contributes to the earlier stages of cognitive impairment in PD.

Blood-based biomarkers have emerged as scalable, minimally invasive, and cost-effective alternatives to PET and CSF measures of AD pathology *in vivo* (13). Among these, plasma phosphorylated tau at threonine 217 (p-tau217) has shown high diagnostic accuracy, outperforming other plasma biomarkers such as plasma phosphorylated tau at threonine 181 (p-tau181) (14). In recent diagnostic guidelines, p-tau217 is now recognized as a highly sensitive and reliable marker of AD pathology (13,15–17). Moreover, there has been particular interest in p-tau217 because elevation of this marker is believed to reflect early tau phosphorylation events that are closely linked to early Aβ accumulation and therefore may detect early AD pathology even before widespread tau aggregation and NFT accumulation has taken place (18).

In contrast to AD, where plasma p-tau217 has been extensively validated, its application in PD remains largely unexplored. To date, most studies have relied on measurement of amyloid β42 and/or p-tau181(19–22). In PD, CSF Aβ42 is generally lower in cognitively impaired PD patients compared to unimpaired PD patients, consistent with patterns observed in AD (23,24). Furthermore, lower Aβ42 concentrations have been associated with increased tau burden and worse baseline cognitive performance across PD groups with or without dementia (11). Interestingly, in cognitively unimpaired PD patients, p-tau181 levels have been found to be lower (i.e., less abnormal) than those of age-matched older adults, suggesting lesser, or possibly delayed, AD pathology compared to what would normally be expected due to age (25–27). Findings regarding the association between p-tau181 levels and cognitive performance in PD patients without dementia have also been inconsistent, further raising questions about the degree to which AD pathology contributes to the early cognitive changes of PD (22). This is in contrast to PDD, where the role of AD pathology, and the value of p-tau181 in particular, is more clearly established (28).

Given the limited and variable findings on fluid biomarkers of AD pathology in relation to the earlier stages of cognitive impairment in PD, the objectives of this study were to determine whether plasma p-tau217 is associated with cognitive impairment or domain-specific cognitive performance on neuropsychological testing in PD without dementia. We also assessed the link between cognition and plasma neurofilament light chain (NfL) and glial fibrillary acidic protein (GFAP), which are non-specific biomarkers of neurodegeneration, with GFAP reflecting astroglial activation and NfL reflecting axonal damage (29–31). Finally, we conducted exploratory analyses to examine the association between plasma biomarkers and other non-motor and motor domains of PD.

## Methods

### Participants

Participants were recruited from the Quebec Parkinson Network (QPN), an ongoing clinical registry of individuals with PD who were referred to the network by their neurologist, as well as healthy controls recruited among friends and relatives of PD participants (32). To ensure a representative PD sample, the only exclusion criteria for enrolment in QPN was major brain lesions such as brain tumor or large stroke. Controls were excluded if they had a first-degree relative with PD or atypical parkinsonism, a diagnosis of a neurological disease, and if they had taken dopamine receptor blockers, metoclopramide, or reserpine in the last 6 months. The participants in QPN have undergone varying degrees of clinical research assessments, which can include Movement Disorders Society Unified Parkinson Disease Rating Scale (MDS-UPDRS) Parts I-III, Montreal Cognitive Assessment (MoCA), and comprehensive neuropsychological testing, in addition to biomarker assessments such as blood sampling. Participants were included in the present analysis if they had available plasma samples for biomarker analysis, were 40 years old or older and had completed either the MoCA or neuropsychological testing within six months from the blood draw. MoCA and neuropsychological assessments were completed during the same visit as blood draws for 86% and 90% of patients respectively. The average interval between cognitive testing and blood sampling was 0.1 months (SD=1.2) for MoCA and 0.1 months (SD=0.8) for neuropsychological assessments. In order to exclude participants with possible PDD, participants who had both a MoCA score <18 and who self-reported cognitive impairment interfering with normal activities (MDS-UPDRS I.I ≥3) were excluded from this analysis (33,34). Among remaining participants, two PD participants had MoCA scores <18, but these participants did not report interference with daily activities, so they were retained in the analyses.

PD participants completed the MDS-UPDRS Parts I (Non-Motor Aspects of Experiences of Daily Living), II (Motor Aspects of Experiences of Daily Living), and III (Motor Examination) and completed all assessments while on their usual dopaminergic medication regimen (35). Testing took place in English or French. The study was approved by the McGill University Health Centre Research Ethics Board and written informed consent was obtained from all participants.

### Processing of blood samples

Blood samples were centrifuged at 4°C for 12 minutes at 250 g to separate the plasma. Aliquots of 250 μL were then transferred into 1 mL cryovials and stored at -80°C for long-term preservation.

### Plasma biomarkers

Plasma concentrations of p-tau217, NfL, and GFAP were quantified using Lumipulse® G assay kits (Fujirebio, Japan) on the fully automated Lumipulse® G1200 platform, which employs chemiluminescent enzyme immunoassay (CLEIA) technology based on monoclonal antibody-coated beads for antigen capture and detection. All plasma samples were previously frozen, then thawed on wet ice (single freeze-thaw cycle only), equilibrated to room temperature for a minimum of 30 minutes, vortexed for 10 seconds, and centrifuged at 2000 g for 5 minutes prior to analysis, in accordance with manufacturer guidelines. Analyses were conducted in batch using a single lot per analyte. P-tau217 was available for 223 participants, whereas GFAP and NfL were measured in a subset of 126 participants.

Plasma p-tau217 positivity was defined using a 0.156 pg/mL cutoff derived from ROC analysis maximizing the Youden index (sensitivity = 0.89, specificity = 0.96) in a cohort of 100 individuals across cognitively unimpaired, mild cognitive impairment (MCI) and dementia stages (Chan T, Rahmouni N, Zheng Y, et al. Validation of plasma p-tau217, plasma p-tau181 and CSF p-tau181 automated assays using positron emission tomography. *Alzheimer’s Dement.* Submitted and under review).

### Measurements of cognition

#### Neuropsychological testing

Participants completed neuropsychological testing as previously described (36). Attention and working memory were measured using the Digit Span (Forward and Backward) subtests from the Wechsler Adult Intelligence Scale Fourth Edition (WAIS-IV) and the Trail Making Test Part A (TMT A). Executive function was measured using the TMT B, the Delis–Kaplan Executive Function System (D-KEFS) Color-Word Interference Test (CWIT; Condition 3: Inhibition time), and the Brixton Spatial Anticipation Test (BSAT; number of errors). Learning and memory was measured using the Hopkins Verbal Learning Test revised (HVLT-R: total from Trials 1-3 and Delayed Recall) and the Rey-Osterrieth Complex Figure Test (RCFT: Immediate and Delayed recalls). Language was measured using the Semantic Verbal Fluency (animals and actions), Letter Verbal Fluency (FAS), and the Boston Naming Test 60 items (BNT-60). Visuospatial function was measured using the Clock Drawing Test (CDT; copy) and the RCFT (copy). Raw scores were converted to z-scores based on the mean and standard deviation of the full sample (PD and controls) and z-scores were averaged within each domain to generate composite scores. TMT A, TMT B, D-KEFS CWIT, and BSAT scores were reverse coded so that higher values reflected better cognitive performance across all assessments. For TMT B, completion times exceeding 300 seconds were capped at 300 seconds, consistent with standard practice that considers this threshold as the poorest possible performance. No participant exceeded 150 seconds on the TMT-A, the standard threshold indicating the poorest possible performance on this task.

#### Classification of cognitive impairment using MoCA

In keeping with previous recommendations, we used a MoCA cutoff score of <26 to identify participants with significant cognitive impairment (34,37).

#### Self-reported cognitive impairment

Item I.1 of the MDS-UPDRS evaluates subjective cognitive impairment reported by the patient (35). We classified patients with a score > 1 as having significant self-reported cognitive impairment (i.e., reporting at least “Clinically evident cognitive dysfunction, but only minimal interference with the ability to carry out normal activities and social interactions” or worse).

### Statistical analyses

Between-group comparisons were performed using t-tests for normally distributed continuous variables and Wilcoxon rank-sum tests otherwise, and chi-square (χ²) tests for categorical variables. For univariate correlations between the different biomarkers, we used Pearson correlation coefficients or Spearman rank correlation coefficients if the assumption of normality was not met (as determined by visual inspection of histograms and Shapiro-Wilk tests). Group differences in the proportion of participants with plasma p-tau217 levels ≥ 0.156 pg/mL were assessed using Pearson’s Chi-square test.

We conducted linear regressions to examine the associations between the different plasma biomarker levels and neuropsychological performance in each cognitive domain; biomarker concentrations were z-scored across the full sample prior to inclusion as predictors in order to allow for comparison of effect sizes. To determine if biomarker levels differed according to cognitive impairment status (i.e. MoCA < 26 and self-reported MDS-UPDRS I.1 > 1) we conducted separate linear regression models for each biomarker. Lastly, we conducted linear regressions to examine associations between plasma biomarkers and MDS-UPDRS (Parts I, II and III). All models controlled for age, sex, education, and disease duration since diagnosis. All p-values were adjusted for multiple comparisons using the false discovery rate (FDR) procedure. Statistical analyses were conducted using R version 4.3.1 (2023-06-16).

## Results

### Participant Characteristics

A total of 195 patients with PD and 70 controls had available blood samples for plasma biomarker analyses. After applying the age exclusion, 185 PD patients and 63 controls remained. Of these, 165 PD patients and 58 controls completed either MoCA or neuropsychological testing. MoCA data were available for 151 PD patients and 55 controls, neuropsychological data for 140 PD patients and 52 controls, and MDS-UPDRS data for 157 PD patients. Demographic and clinical characteristics are summarized in Table 1. The proportion of males was significantly higher in the PD group than in controls. As expected, MoCA scores were significantly lower in the PD group compared to the controls.

**Table 1.**
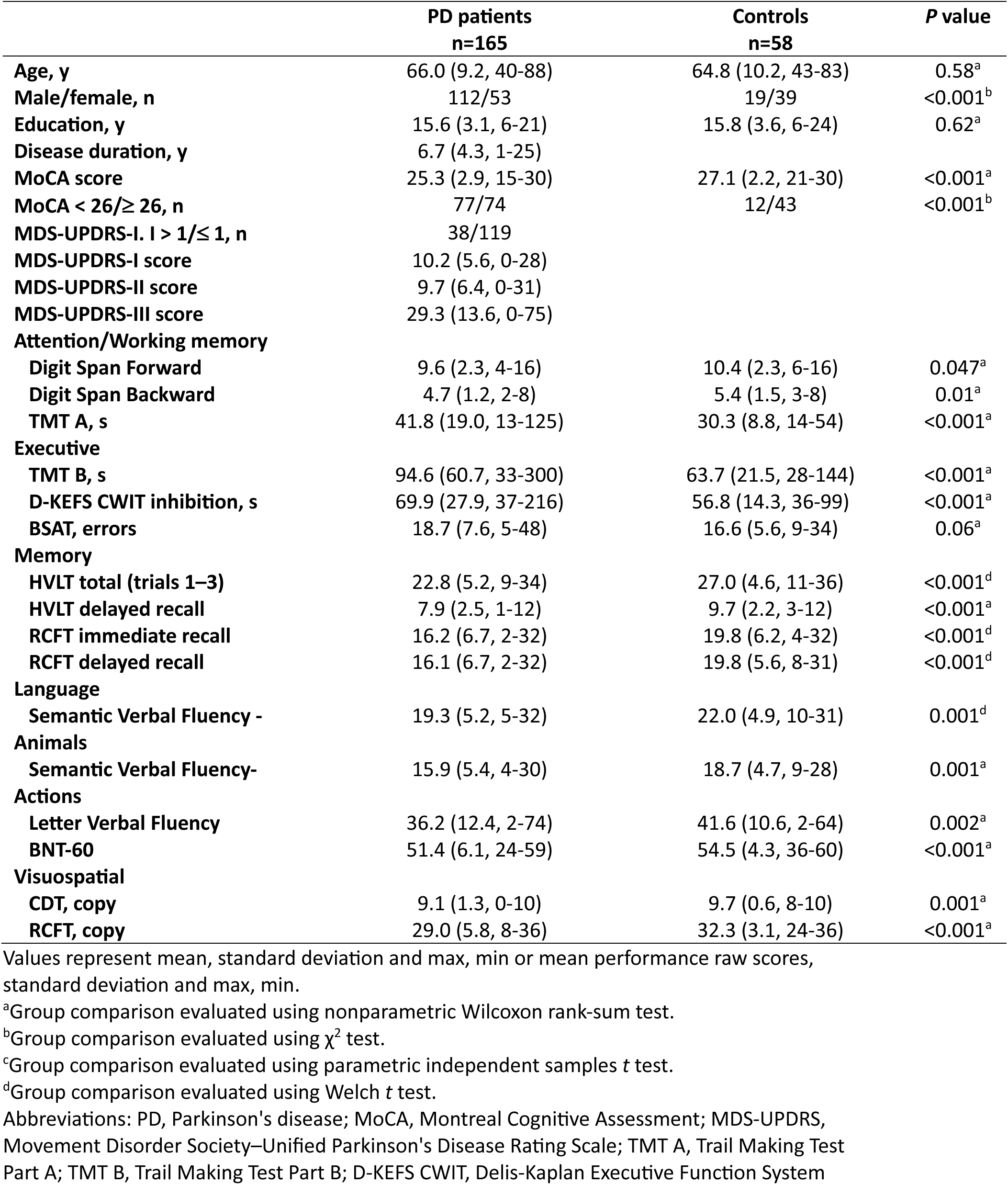
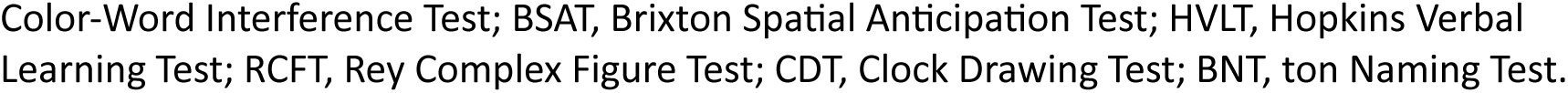
Sample Characteristics.

|  | PD patients<br>n=165 | Controls<br>n=58 | P value |
| --- | --- | --- | --- |
| Age, y | 66.0 (9.2, 40-88) | 64.8 (10.2, 43-83) | 0.58<sup>a</sup> |
| Male/female, n | 112/53 | 19/39 | <0.001<sup>b</sup> |
| Education, y | 15.6 (3.1, 6-21) | 15.8 (3.6, 6-24) | 0.62<sup>a</sup> |
| Disease duration, y | 6.7 (4.3, 1-25) |  |  |
| MoCA score | 25.3 (2.9, 15-30) | 27.1 (2.2, 21-30) | <0.001<sup>a</sup> |
| MoCA < 26/≥ 26, n | 77/74 | 12/43 | <0.001<sup>b</sup> |
| MDS-UPDRS-I. I > 1/≤ 1, n | 38/119 |  |  |
| MDS-UPDRS-I score | 10.2 (5.6, 0-28) |  |  |
| MDS-UPDRS-II score | 9.7 (6.4, 0-31) |  |  |
| MDS-UPDRS-III score | 29.3 (13.6, 0-75) |  |  |
| Attention/Working memory |  |  |  |
| Digit Span Forward | 9.6 (2.3, 4-16) | 10.4 (2.3, 6-16) | 0.047<sup>a</sup> |
| Digit Span Backward | 4.7 (1.2, 2-8) | 5.4 (1.5, 3-8) | 0.01<sup>a</sup> |
| TMT A, s | 41.8 (19.0, 13-125) | 30.3 (8.8, 14-54) | <0.001<sup>a</sup> |
| Executive |  |  |  |
| TMT B, s | 94.6 (60.7, 33-300) | 63.7 (21.5, 28-144) | <0.001<sup>a</sup> |
| D-KEFS CWIT inhibition, s | 69.9 (27.9, 37-216) | 56.8 (14.3, 36-99) | <0.001<sup>a</sup> |
| BSAT, errors | 18.7 (7.6, 5-48) | 16.6 (5.6, 9-34) | 0.06<sup>a</sup> |
| Memory |  |  |  |
| HVLT total (trials 1–3) | 22.8 (5.2, 9-34) | 27.0 (4.6, 11-36) | <0.001<sup>d</sup> |
| HVLT delayed recall | 7.9 (2.5, 1-12) | 9.7 (2.2, 3-12) | <0.001<sup>a</sup> |
| RCFT immediate recall | 16.2 (6.7, 2-32) | 19.8 (6.2, 4-32) | <0.001<sup>d</sup> |
| RCFT delayed recall | 16.1 (6.7, 2-32) | 19.8 (5.6, 8-31) | <0.001<sup>d</sup> |
| Language |  |  |  |
| Semantic Verbal Fluency - Animals | 19.3 (5.2, 5-32) | 22.0 (4.9, 10-31) | 0.001<sup>d</sup> |
| Semantic Verbal Fluency-Actions | 15.9 (5.4, 4-30) | 18.7 (4.7, 9-28) | 0.001<sup>a</sup> |
| Letter Verbal Fluency | 36.2 (12.4, 2-74) | 41.6 (10.6, 2-64) | 0.002<sup>a</sup> |
| BNT-60 | 51.4 (6.1, 24-59) | 54.5 (4.3, 36-60) | <0.001<sup>a</sup> |
| Visuospatial |  |  |  |
| CDT, copy | 9.1 (1.3, 0-10) | 9.7 (0.6, 8-10) | 0.001<sup>a</sup> |
| RCFT, copy | 29.0 (5.8, 8-36) | 32.3 (3.1, 24-36) | <0.001<sup>a</sup> |

Values represent mean, standard deviation and max, min or mean performance raw scores, standard deviation and max, min.
<sup>a</sup>Group comparison evaluated using nonparametric Wilcoxon rank-sum test.
<sup>b</sup>Group comparison evaluated using $\chi^2$ test.
<sup>c</sup>Group comparison evaluated using parametric independent samples *t* test.
<sup>d</sup>Group comparison evaluated using Welch *t* test.
Abbreviations: PD, Parkinson's disease; MoCA, Montreal Cognitive Assessment; MDS-UPDRS, Movement Disorder Society–Unified Parkinson's Disease Rating Scale; TMT A, Trail Making Test Part A; TMT B, Trail Making Test Part B; D-KEFS CWIT, Delis-Kaplan Executive Function System
Color-Word Interference Test; BSAT, Brixton Spatial Anticipation Test; HVL, Hopkins Verbal Learning Test; RCFT, Rey Complex Figure Test; CDT, Clock Drawing Test; BNT, ton Naming Test.

Pairwise correlations between plasma biomarkers were examined in the PD group: as expected all biomarkers were positively correlated with one another, with the strongest association observed between p-tau217 and GFAP (r_ptau217-GFAP_ = 0.45; *p_FDR_* < 0.001), followed by GFAP and NfL (r_GFAP-NFL_ = 0.35; *p_FDR_* < 0.001) and NfL and p-tau217 (r_NFL-ptau217_= 0.33; *p_FDR_* < 0.001).

### Group Differences in Plasma Biomarker Levels

PD patients and controls did not differ significantly after correction for multiple comparisons with respect to adjusted p-tau217 (*p_FDR_* = 0.865), GFAP (*p_FDR_* = 0.076), and NfL (*p_FDR_* = 0.123), although mean GFAP and NFL levels were numerically higher in PD (Figure 1, Supplementary Table 1).

**Figure 1.**
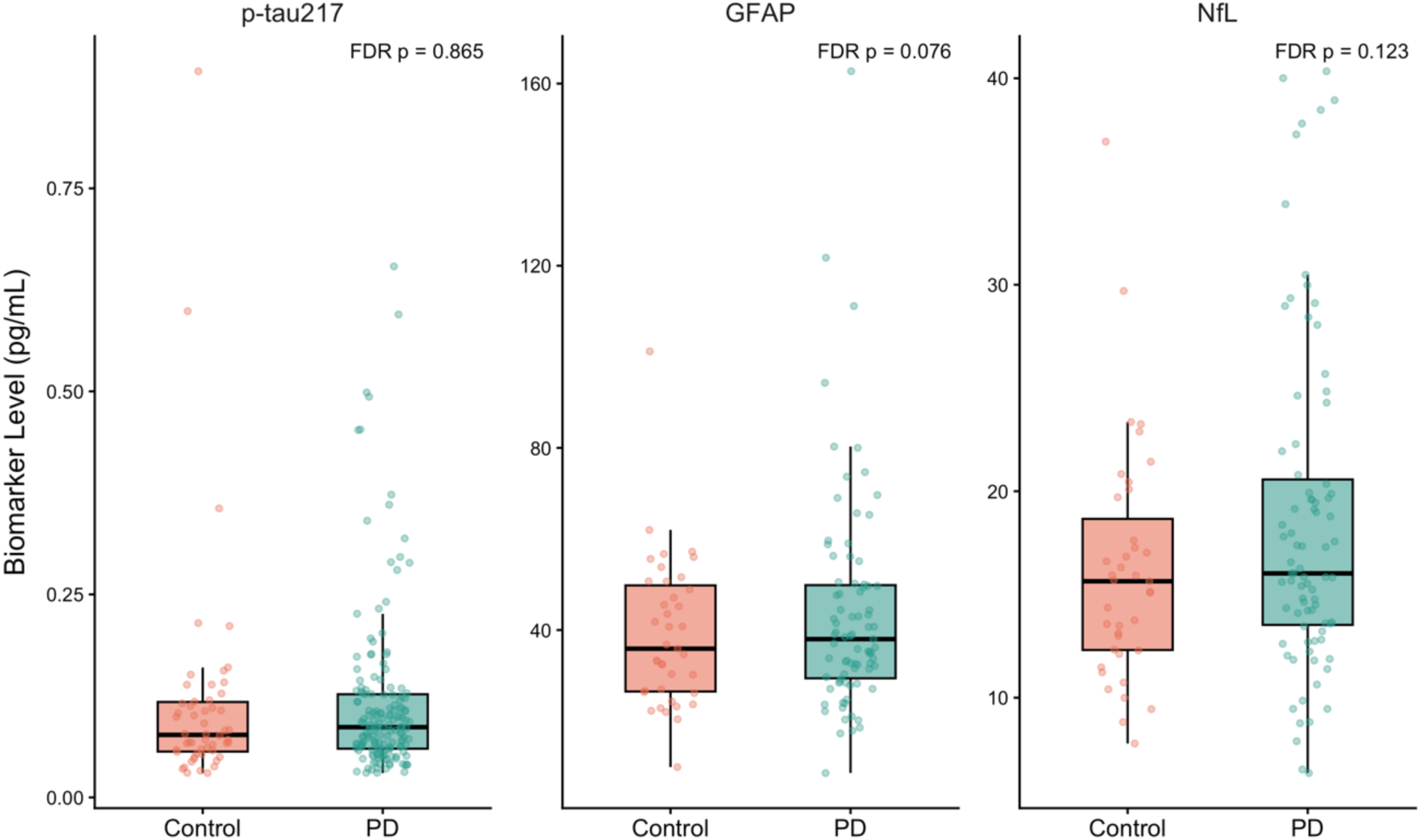
Plasma biomarker levels by diagnostic group. Boxplots of raw plasma p-tau217, GFAP, and NfL concentrations (pg/mL) in PD patients and controls. Boxplots display the median (horizontal line), interquartile range (box limits), and data spread within 1.5 times the IQR (whiskers). Individual dots represent raw data points, jittered for visibility. FDR-corrected p- values are based on linear regression models adjusted for age at blood draw, sex and years of education.

### Association between plasma biomarkers and neuropsychological performance in PD

In the PD group, higher plasma p-tau217 was associated with poorer visuospatial function (β = -0.18, 95% CI = -0.36 to -0.01, *p* = 0.04, *p_FDR_* = 0.20) and higher GFAP was associated with poorer memory (β = -0.21, 95% CI = -0.40 to -0.02, *p* = 0.03, *p_FDR_* = 0.20), however, neither association remained significant after FDR correction. None of the other cognitive domains were significantly associated with plasma p-tau217, GFAP or NfL levels (Table 2). Unadjusted pairwise correlations between biomarker levels and cognitive domain performance are presented in Supplementary Figure 1.

**Table 2.** Associations Between Plasma Biomarkers and Neuropsychological Performance in PD.

| Cognitive Domain | p-tau217 Beta (95% CI) | p (p<sub>FDR</sub>) | GFAP Beta (95% CI) | p (p<sub>FDR</sub>) | NfL Beta (95% CI) | p (p<sub>FDR</sub>) |
| --- | --- | --- | --- | --- | --- | --- |
| Attention/WM | -0.08 (-0.22 to 0.06) | 0.25 (0.37) | -0.17 (-0.37 to 0.02) | 0.08 (0.20) | -0.10 (-0.30 to 0.09) | 0.30 (0.37) |
| Executive | -0.10 (-0.28 to 0.08) | 0.29 (0.37) | -0.25 (-0.49 to -0.004) | 0.05 (0.20) | -0.21 (-0.45 to 0.03) | 0.09 (0.20) |
| Memory | -0.12 (-0.25 to 0.02) | 0.09 (0.20) | -0.21 (-0.40 to -0.02) | 0.03 (0.20) | -0.09 (-0.28 to 0.11) | 0.37 (0.42) |
| Language | -0.04 (-0.19 to 0.12) | 0.65 (0.65) | -0.14 (-0.36 to 0.09) | 0.23 (0.37) | -0.08 (-0.31 to 0.15) | 0.48 (0.51) |
| Visuospatial | -0.18 (-0.36 to -0.01) | 0.04 (0.20) | -0.21 (-0.46 to 0.04) | 0.10 (0.20) | -0.17 (-0.42 to 0.08) | 0.18 (0.33) |

Table presents the beta estimates derived from linear regressions examining the association between each of the plasma biomarkers and performance on each of the neuropsychological cognitive domains. All models were adjusted for age, sex, education and disease duration. Biomarker values were standardized to allow for comparison of beta estimates across models.
Abbreviations: WM, Working Memory.

When examining the association between p-tau217 positivity vs negativity and neuropsychological performance, p-tau217 positivity was associated with poorer visuospatial function (β = -0.52, 95% CI = -0.94 to -0.10, *p* = 0.02; Supplementary Table 2), but this association did not remain significant after FDR (*p_FDR_* = 0.08). P-tau217 positivity was not significantly associated with any other cognitive domain (*p_FDR_* > 0.15).

### Association between plasma biomarkers and cognitive categorization

No significant differences in plasma biomarker levels were observed between PD patients stratified based on MoCA score (*p_FDR_* > 0.31; Table 3). In patients with self-reported cognitive impairment, p-tau217 levels were higher (*p* = 0.03), but this group difference did not remain significant after FDR correction (*p_FDR_* = 0.08; Table 3). No significant group difference was observed for GFAP and NfL.

**Table 3.** Biomarker levels in PD patients with and without cognitive impairment as determined by MoCA score and by self-report.

| Biomarker | PD with MoCA < 26 | PD with MoCA ≥ 26 | *p* (*p*<sub>FDR</sub>) | PD with MDS-UPDRS I.1 ≤ 1 | PD with MDS-UPDRS I.1 > 1 | *p* (*p*<sub>FDR</sub>) |
| --- | --- | --- | --- | --- | --- | --- |
| **P-tau217** | 0.118 (0.094) | 0.125(0.124) | 0.10 (0.31) | 0.108 (0.091) | 0.160 (0.152) | 0.03 (0.08) |
| **GFAP** | 50.6 (24.7) | 38.6 (25.6) | 0.47 (0.47) | 43.07 (27.96) | 48.0 (19.8) | 0.90 (0.90) |
| **NfL** | 20.2 (8.8) | 18.0 (7.7) | 0.37 (0.47) | 18.11 (8.1) | 22.9 (9.5) | 0.17 (0.26) |

Table presents means (SD) of raw biomarker concentrations. Adjusted p-values are derived from linear regressions controlling for age, sex, education, and disease duration. FDR-adjusted p-values use Benjamini-Hochberg across biomarkers.

### Associations between plasma biomarkers and MDS-UPDRS I, II and III in PD

We also examined whether plasma biomarkers of neurodegeneration were associated with other clinical domains in PD (Table 4) using total scores from each MDS-UPDRS subscale. No significant associations were observed for p-tau217 nor GFAP with any subscale of MDS-UPDRS. However, higher NfL levels were significantly associated with worse Part I scores (β = 0.35, 95% CI = 0.10 to 0.60, *p_FDR_* = 0.03) and worse Part II (β = 0.39, 95% CI = 0.17 to 0.61, *p_FDR_* = 0.006) scores, but not with Part III scores (*p_FDR_* = 0.20).

**Table 4.** Associations Between Plasma Biomarkers and functional and motor outcomes in PD.

|  | p-tau217 Beta (95% CI) | *p* (*p*<sub>FDR</sub>) | GFAP Beta (95% CI) | *p* (*p*<sub>FDR</sub>) | NfL Beta (95% CI) | *p* (*p*<sub>FDR</sub>) |
| --- | --- | --- | --- | --- | --- | --- |
| **Non-Motor Aspects of Daily Living (MDS-UPDRS PART I)** | 0.06 (-0.13 to 0.25) | 0.51 (0.65) | 0.12 (-0.13 to 0.38) | 0.34 (0.62) | 0.35 (0.10 to 0.60) | 0.01 (0.03) |
| **Motor Aspects of Daily Living (MDS-UPDRS PART II)** | 0.04 (-0.13 to 0.22) | 0.62 (0.70) | 0.01 (-0.22 to 0.24) | 0.91 (0.91) | 0.39 (0.17 to 0.61) | 0.001 (0.006) |
| **Motor Examination (MDS-UPDRS PART III)** | 0.10 (-0.07 to 0.27) | 0.26 (0.59) | -0.10 (-0.34 to 0.14) | 0.43 (0.64) | 0.23 (-0.02 to 0.47) | 0.07 (0.20) |

Table presents the beta estimates derived from linear regressions examining the association between each of the plasma biomarkers and scores on MDS-UPDRS Parts I, II and III. All models adjusted for age, sex, education and disease duration. Biomarker values were standardized to allow for comparison of beta estimates across models.

## Discussion

An important gap in PD research is our limited understanding of the role of AD pathology in the earlier stages of cognitive impairment. This study, conducted using the QPN patient registry, examined whether plasma p-tau217, which is considered to be the most sensitive fluid biomarker for the early presence of AD pathology (14,38), is associated with cognitive impairment and domain-specific cognitive performance in PD. We identified several key findings. First, p-tau217 levels were not higher in the individuals with PD compared to controls, despite the generally worse cognitive function in the PD group. Second, contrary to our predictions, plasma p-tau217 levels were not significantly associated with neuropsychological performance, nor with bedside cognitive testing and self-reported cognitive impairment. We found a similar pattern for plasma GFAP and NfL. These findings suggest that AD co-pathology may not be a major contributor to the earlier stages of cognitive impairment in PD.

In our sample of PD patients without dementia, we found only equivocal or absent relationships between p-tau217 and cognitive impairment, both when examining performance-based and self-reported measures of cognition. Our findings are broadly consistent with several recent studies relying on p-tau181, which have shown that p-tau181 level is not increased in PD patients without dementia (39) and that p-tau181 levels are not correlated with cognitive performance (40). Interestingly, studies reporting on Aβ42 in PD patients in the earlier stage of disease show a somewhat different pattern in that levels of Aβ42 are already lower in early PD compared to controls, and low Aβ42 levels have been more consistently associated with cognitive impairment in early PD, suggesting a divergence between Aβ42 levels and p-tau levels that contrasts with the pattern seen in AD (25,41,42).

The absence of clear links between p-tau217 and cognition in PD is in keeping with the long-standing dual syndrome of dementia hypothesis for PD, which posits that early cognitive changes are mediated by dopaminergic loss (43,44) and other subcortical neuromodulatory systems (36). In contrast, the more advanced, and generally later, state of dementia is believed to be due to more prominent cortical involvement, either due to the progression of PD pathology, or additionally due to co-morbid AD pathology (45,46). Indeed, in our sample, though not statistically significant, the largest effect sizes for the associations between p-tau217 and cognitive performance were observed for the domains of memory and visuospatial function, which are also the hallmarks of early AD-associated cognitive impairment (1,47,48). Together, these findings raise the possibility that p-tau217 may show stronger associations with cognition in PD patients with more advanced cognitive or functional impairment, a stage more likely to represent the combined effects of PD and AD pathology. Studies simultaneously assessing amyloid and p-tau217 markers will be necessary to confirm whether the weak association between p-tau217 and cognition in our sample reflects a true absence of amyloid pathology in early cognitive loss in PD. PET studies of amyloid deposition will help validate the use of p-tau217 as a marker of AD pathology in PD and to determine whether there is a relationship between the spatial distribution of amyloid and tau and the cognitive phenotype in early PD.

It is also interesting to consider our results in light of results from a recent study showing that in individuals with idiopathic/isolated REM sleep behavior disorder, p-tau181 was a strong predictor of later conversion to dementia with Lewy bodies (DLB) (49). This suggests that patients who develop motor-onset PD instead of dementia-onset DLB may have effectively been “filtered out” from having significant AD pathology, i.e. they may be the individuals, who owing to genetics, environmental, or other factors, have lower AD pathology and thus manifest with a motor-predominant phenotype. Indeed, amyloid-β (Aβ) pathology is relatively common in older adults with approximately 20% of individuals around age 65 demonstrating a positive Aβ PET scan and with this prevalence increasing further with age (50,51). Prodromal Lewy body disease progresses slowly, with an average interval of 13 years between symptom onset of iRBD and conversion to PD or DLB (52–54), and over 20 years between onset of hyposmia and conversion (55,56). During this prolonged prodromal period, any co-occurring AD pathology (even at relatively low levels) may compound with Lewy body pathology, precipitating a primary dementia phenotype (i.e., DLB), whereas individuals with AD pathology in the lower range might be more likely to present with a motor-predominant phenotype (i.e, PD). In this sense, DLB represents a competing risk for motor-onset PD. Over time, however, age-related amyloid accumulation in individuals with a motor-onset phenotype continues such that at advanced stages of PD, accumulated amyloid pathology increasingly contributes to the clinical manifestations of PD, in particular dementia.

Neither GFAP nor NFL plasma levels were significantly associated with neuropsychological performance or classification of cognitive impairment in the PD patients. Previous findings linking GFAP and NfL with cognition in PD are heterogeneous (39,57,58). Generally, however, in PD, GFAP and NfL appear to be most elevated in patients with dementia, even when comparing groups with comparable disease duration (39,59). This suggests that while these biomarkers are not specific to either AD or PD, increased levels may reflect overall neurodegeneration, rather than pathology specific to AD or PD (39). In our cohort, GFAP and NfL levels also did not differ between PD and controls. Previous studies comparing these biomarkers in PD and controls have yielded somewhat conflicting results, though generally, when comparing early or cognitively unimpaired PD patients with older adults, differences are either small or not significant, and might simply reflect the characteristics of the control group used for comparison, which varies across studies depending on the degree to which strict inclusion criteria were applied to the control group (40). For the present study, no exclusion criteria other than age were applied to the control group in order to obtain a representative sample, and as a result it is possible that this sample would include individuals with early sub-clinical neurodegeneration.

In addition to cognitive outcomes, we examined associations between plasma biomarkers and other PD symptom domains. Neither p-tau217 nor GFAP showed significant associations with non-motor or motor features. However, higher NfL levels were associated with greater impairments in non-motor and motor aspects of daily living as captured by the MDS-UPDRS parts I and II, although not with the motor examination scores. These findings indicate that while NfL may not be specifically sensitive to early cognitive impairment, it may nevertheless reflect the severity of the neurodegenerative state in PD. This is consistent with prior reports of its utility as a marker of motor and non-motor symptoms in early PD (42).

Our study has several limitations. First, we were unable to assess amyloid-β 42/40 status in our sample. Although p-tau217 is a sensitive marker of AD pathology in AD cohorts, its ability to detect the presence of AD pathology in PD remains uncertain. Amyloid-β and tau PET imaging would have also provided additional insight into how p-tau217 trajectories in PD may relate to the spatial accumulation of amyloid-β and tau. Longitudinal studies incorporating fluid and PET imaging-derived markers of both tau and amyloid-β accumulation will also be necessary to determine the causal link between AD pathology and cognitive impairment in PD, and to disentangle these effects from PD-specific mechanisms of neurodegeneration.

In conclusion, our study found that plasma p-tau217 was not clearly associated with cognitive impairment in PD. This lack of association may reflect either a limited burden of AD co-pathology in our cohort or a limited sensitivity of p-tau217 to detect such pathology in the context of PD. Early cognitive impairment in PD may primarily arise from PD pathology-related mechanisms, with the effects of co-morbid pathology being more evident only in PD patients who develop dementia. Overall, these findings suggest that p-tau217 alone is unlikely to capture mild cognitive impairment in PD underscoring the need to consider both AD-related and PD-specific processes when investigating determinants of cognitive impairment in PD.

## Data Availability

All data produced in the present study are available upon reasonable request to the authors.

## Acknowledgements

We thank the Quebec Parkinson Network and all our participants for their contribution.

## Author contributions

TK: conception, analysis, writing and revising the manuscript; TC: biomarker analysis, revising the manuscript; SS: analysis, revising the manuscript; AD: conception, revising the manuscript; PRN: funding, biomarker analysis, revising the manuscript; JFG: conception, funding, revising the manuscript; AD: conception, funding, revising the manuscript; RBP: conception, supervision, funding, writing and revising the manuscript; MS: conception, supervision, funding, writing and revising the manuscript.

