## Supplemental Figure 1 and Tables for "Association between plasma phosphorylated tau-217 and cognition in Parkinson’s disease"

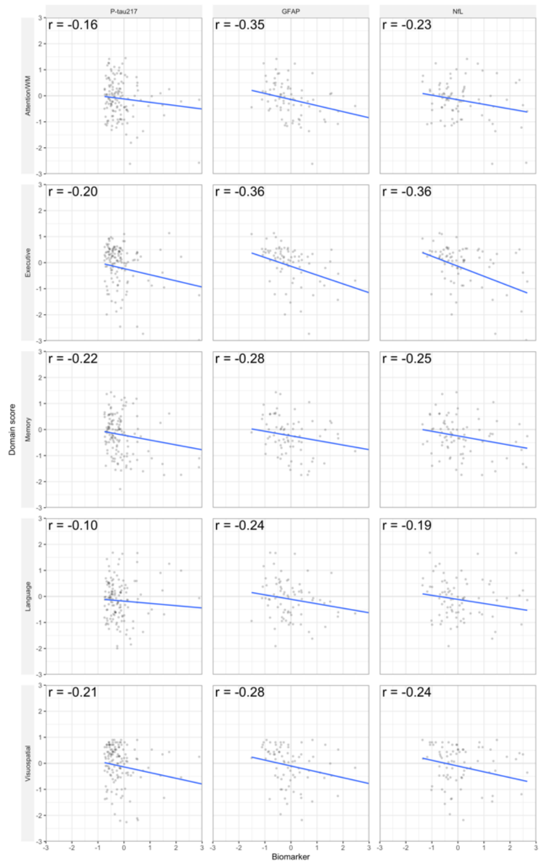


**Supplementary Figure 1. Pairwise correlations between biomarker levels and cognitive domain scores.** Both biomarker levels and cognitive domain scores were standardized (z-scores). Points represent individuals; lines are least-squares fits. Each panel shows Pearson's r coefficient.

**Supplementary Table 1.** Descriptive statistics and adjusted group comparisons for plasma biomarkers

| **Plasma Biomarkers** | | **Controls n=58** | **PD n=165** | ***p*** **(*$p_{FDR}$*)** |
|---|---|---|---|---|
| **P-tau217** | Mean (SD) | 0.122 (0.146) | 0.116 (0.104) | 0.865 (0.865) |
| | Min/Max | 0.030/0.894 | 0.030/0.654 | |
| **GFAP[a]** | Mean (SD) | 37.8 (16.9) | 43.6 (23.7) | 0.025 (0.076) |
| | Min/Max | 9.9/101.2 | 8.6/162.7 | |
| **NfL[a]** | Mean (SD) | 15.8 (5.9) | 18.6 (8.1) | 0.082 (0.123) |
| | Min/Max | 6.4/36.9 | 6.4/40.3 | |

[a]GFAP anf NfL levels were only available for 85 PD patients and 41 controls.

When assessing the proportion of individuals above the p-tau217 positivity cutoff, 17.3 % of PD patients and 12.9% of controls were above the cutoff. However, this difference was not statistically significant (p = 0.46).

**Supplementary Table 2.** Associations Between Plasma p-tau217 positivity and Neuropsychological Performance in PD

| Cognitive Domain | p-tau217 positivity Beta (95% CI) | $p$ ($p_{FDR}$) |
|---|---|---|
| **Attention/WM** | -0.26 (-0.60 to 0.07) | 0.12 (0.21) |
| **Executive** | -0.42 (-0.86 to 0.02) | 0.06 (0.15) |
| **Memory** | -0.20 (-0.53 to 0.13) | 0.23 (0.29) |
| **Language** | 0.13 (-0.25 to 0.51) | 0.49 (0.49) |
| **Visuospatial** | -0.52 (-0.94 to -0.10) | 0.02 (0.08) |

Table presents the beta estimates derived from linear regressions examining the association between each of the plasma p-tau217 positivity (p-tau217 levels ≥0.156) vs negativity (p-tau217 levels <0.156) and performance on each of the neuropsychological cognitive domains. All models were adjusted for age, sex, education and disease duration. Abbreviation: WM, Working Memory.
